# A Systematic Review: Cardiovascular effects of Systemic Sclerosis

**DOI:** 10.1101/2023.10.20.23297312

**Authors:** Dedeepya Panchumarthi, Sasidhar Reddy Bathula, Sai Tapasvi madam, Raghu Ram Shanmukh Nemani, Binay Kumar Panjiyar

**Affiliations:** HARVARD MEDICAL SCHOOL; G.S.L medical college; G.S.L MEDICAL COLLEGE; GSL Medical College

**Keywords:** Cardiology, Rheumatology, Internal medicine, cardiovascular effects, systemic sclerosis, autoimmune diseases, arrhythmias, myocardial infarction, coronary artery diseases, valvular diseases

## Abstract

Systemic sclerosis is a connective autoimmune disorder characterized by the deposition of an extracellular matrix in the internal organs leading to fibrosis and damage to internal organs. Cardiovascular effects of systemic sclerosis include arrhythmias, coronary artery disease, valvular diseases, pericardial diseases, atherosclerosis, myocardial diseases. Thus it also has a fatal effect on the cardiovascular system making it significant to know its epidemiology, pathophysiology, screening techniques, and investigations which help in early diagnosis of the cardiac lesions of systemic sclerosis. Cardiovascular effects of systemic sclerosis is a multidisciplinary approach by a rheumatologist, cardiologist and radiologist. We reviewed 182 articles from renowned journals taken from google scholar and pubmed published between 2010 and 2023 of which 8 articles are focused for in depth review. This systematic review summarizes the effects of systemic sclerosis on cardiovascular system ranging from arrhythmias to pericardial diseases in brief including the pathogenesis, investigations and epidemiology.

## Introduction and Background

Systemic sclerosis a connective autoimmune disorder affecting the skin, internal organs and vasculature resulting in inflammation and fibrosis of the tissues, typically characterized by Raynaud’s phenomenon, digital ischemia, sclerodactyly, cardiac, renal, lung and gut diseases. It is usually seen in females > males predominantly in the age of fourth and fifth decades. Systemic sclerosis is further divided into diffuse and limited cutaneous systemic sclerosis [1] of which limited systemic sclerosis accounts for 70% of cases.

Severity of internal organ involvement is an important prognostic factor for overall survival of which cardiopulmonary manifestations have the worst prognosis. These patients have a five year survival rate as high as 75%.[2]. Cardiac manifestations of systemic sclerosis range from pericardial disease to cardiac failure and death resulting from the various grave presentations that affect every structure of the heart and resulting in pericarditis, arrhythmias, conduction system abnormalities, myocardial disease like myositis, fibrosis of myocardium, coronary artery disease, rarely primary valvular impairment and cardiac failure leading to death. [3]

The exact mechanism for elevated risk of atherosclerotic cardiovascular disease in patients with systemic sclerosis remains incompletely understood. Activation of T-cell and formation of auto antibodies indicates inflammation making it one of the potential mechanisms for development of atherosclerotic disease. In patients with systemic sclerosis an increase in pro inflammatory cytokines including Interleukin (IL-1), IL-6 and C reactive protein hs been observed.[6]

The other possible mechanism can be fibrosis and there is increasing evidence that strongly indicates the involvement of myocardium leads to myocardial fibrosis with inevitable lesions resulting from repeated ischemic foci. Patients from SSc may experience both primary microvascular disease and secondary macrovascular complications such as Ischemic heart disease or pulmonary vascular disease at the same time [7].This systematic review summarizes the cardiac related manifestations in systemic sclerosis its pathophysiology, epidemiology, investigations for diagnosis of the lesions.

## Results

### Methods

This review focuses on clinical studies concerning the effects of systemic sclerosis on the cardiovascular system. We excluded animal studies and publications that only discussed the effects of systemic sclerosis on the cardiovascular system without presenting clinical data. The review follows the guidelines for Preferred Reporting Items for Systematic Reviews and Meta-Analyses (PRISMA) for 2020 in Figure 2 and only uses data collected from published papers, eliminating the need for ethical approval.

**FIGURE 2:**
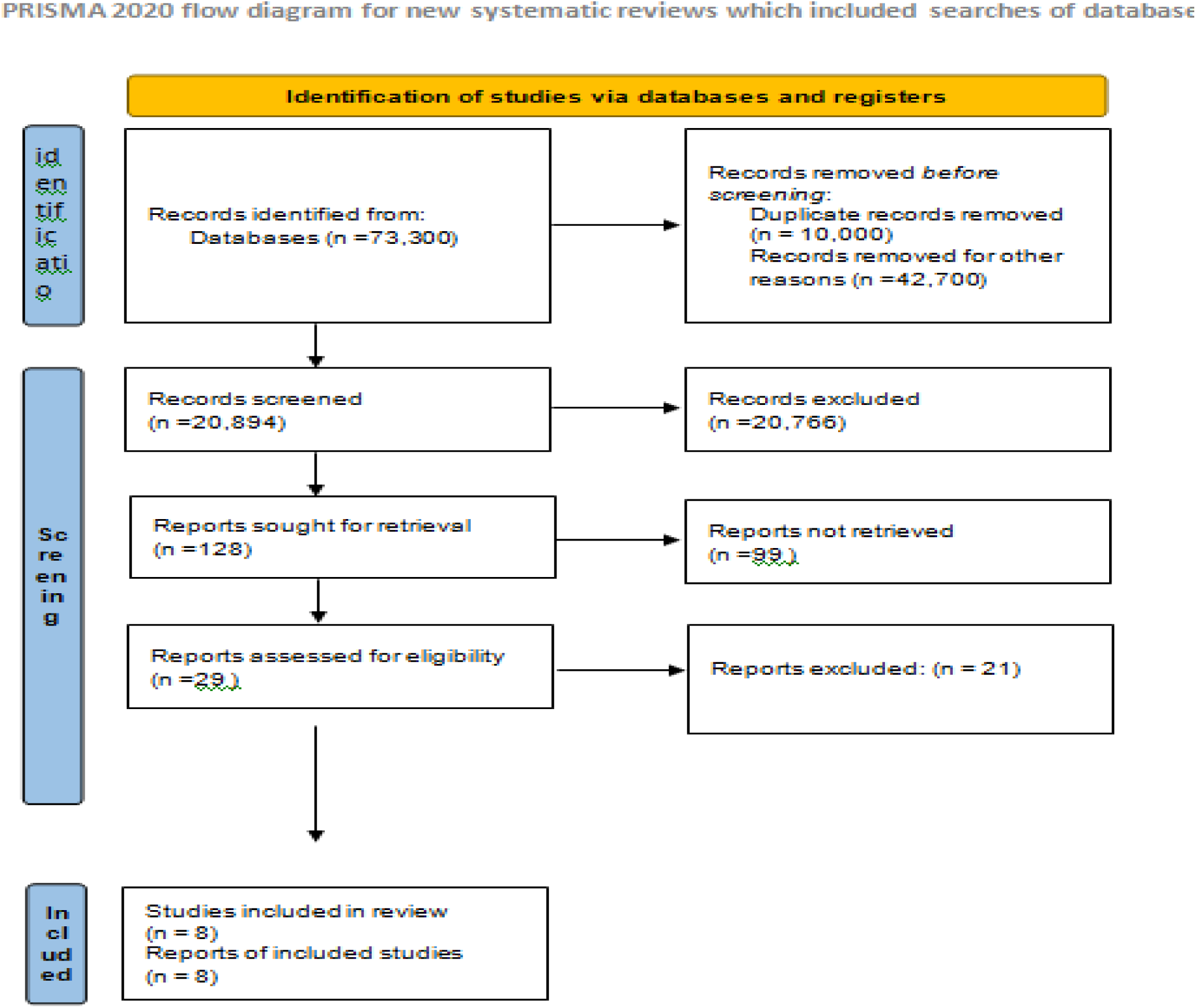
PRISMA flow diagram illustrating the search strategy and study selection process for the systematic review.

The sources used for this review includes Google scholar, PMC free articles, Medline, and PubMed were systematically searched from 2010 to 2023, without language or geographic restrictions. We used different combinations of terms “systemic sclerosis” AND “cardiovascular disease” AND “cardiovascular effects of systemic sclerosis”.

The PRISMA study flow diagram is shown in Figure 1. The systematic literature search of articles published from 2010 to 2023, 20,984 identified articles, of which 20,766 were deemed ineligible based on the titles and/or abstracts. After careful examination of the full text, 8 articles that met the inclusion criteria were identified.

### Inclusion and Exclusion Criteria

We established specific criteria for including and excluding participants to achieve our study goals. Our Criteria can be summarized in Table 1.

**TABLE 1:** Showing criteria adopted during literature selection process.

| **Inclusion criteria** | **Exclusion criteria** |
| --- | --- |
| Human studies | Animal studies |
| 2010 - 2023 | Below 2010 |
| English text | Non english text |
| Gender- all |  |
| Free articles | Papers that need to be Purchased |

### Search Strategy

The population, intervention/condition, control/comparison, and outcome (PICO) criteria were utilized to conduct a thorough literature review. The search was conducted on databases such as PUBMED (including Medline) and Google Scholar Libraries, using relevant keywords, such as cardiovascular effects and systemic sclerosis. The medical subject heading (MeSH) approach for PubMed (including Medline) and Google Scholar as detailed in Table 2, was employed to develop a comprehensive search strategy.

**Table 2:** Showing search strategy, search engines and the search results displayed.

|  | **Databas**<br>**e** | **Search strategy** | **Search**<br>**results** |
| --- | --- | --- | --- |
| 1 | Google<br>scholar | Cardiovascular effects of systemic sclerosis | 16,600 |
| 2 | Pubmed | Cardiovascular effects or systemic sclerosis | 90 |
|  |  | Cardiovascular effects of systemic sclerosis<br>( filtered with year starting from 2010 to 2023 and with article type ) | 4294 |

### Quality appraisal tools

To ensure the reliability of our chosen papers, we utilized various quality assessment tools. We employed the PRISMA checklist and Cochrane bias tool assessment for randomized clinical trials for systematic reviews and meta-analyses. Non-randomized clinical trials were evaluated using the Newcastle-Ottawa tool scale.We assessed the quality of qualitative studies, as shown in Table 3, using the critical appraisal skills program (CASP) checklist. To avoid any confusion in the classification, we utilized the scale for the assessment of narrative review articles (SANRA) to evaluate the article’s quality

**TABLE 3:** Showing quality appraisal tools used.

| **Quality appraisal tools** | **Type of studies** |
| --- | --- |
| Cochrane bias tool assessment | Randomized control trials |
| Newcastle - ottawa tool | Non RCT and observational studies |
| PRISMA Checklist | Systematic reviews |
| SANRA Checklist | Any other without clear method section |

PRISMA: Preferred reporting items for systematic reviews and meta-analyses; SANRA: Scale for the assessment of non-systematic review articles

## Results

After searching through three selected databases, PubMed, Medline, and Google Scholar, we extracted 73,300 articles. We then carefully reviewed each paper and applied specific criteria, which led to excluding 42,700 articles. From the remaining 20,894 papers, we chose 128 eliminating duplicates or unsatisfactory titles and abstracts. We closely examined 128 papers and excluded 120 more as their content did not meet our inclusion criteria. Finally, we conducted a thorough quality check on the remaining eight papers, which all met our criteria. These eight articles are included in our final systematic review. Table 4 provides a detailed description of each.

**TABLE 4:**
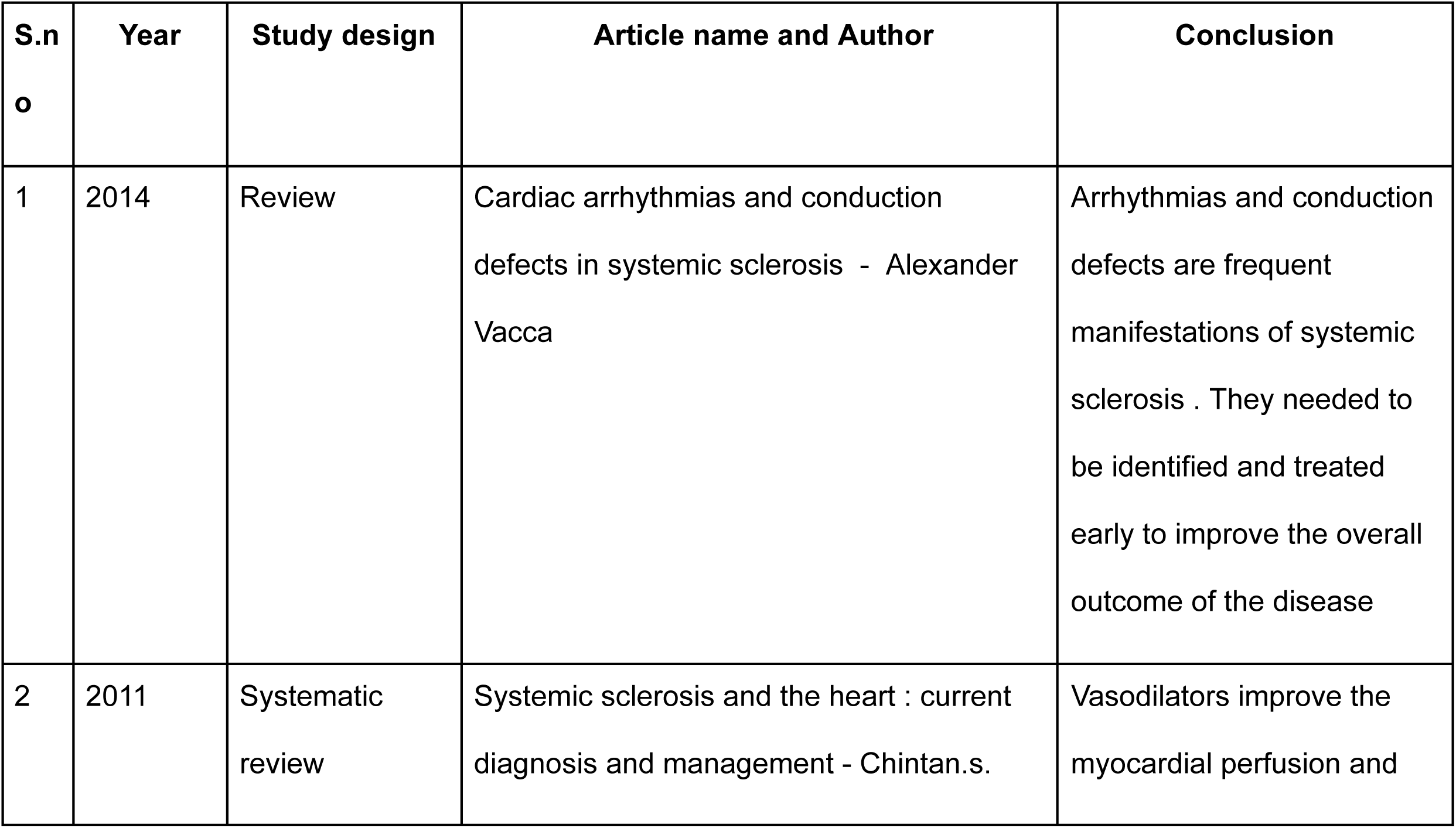
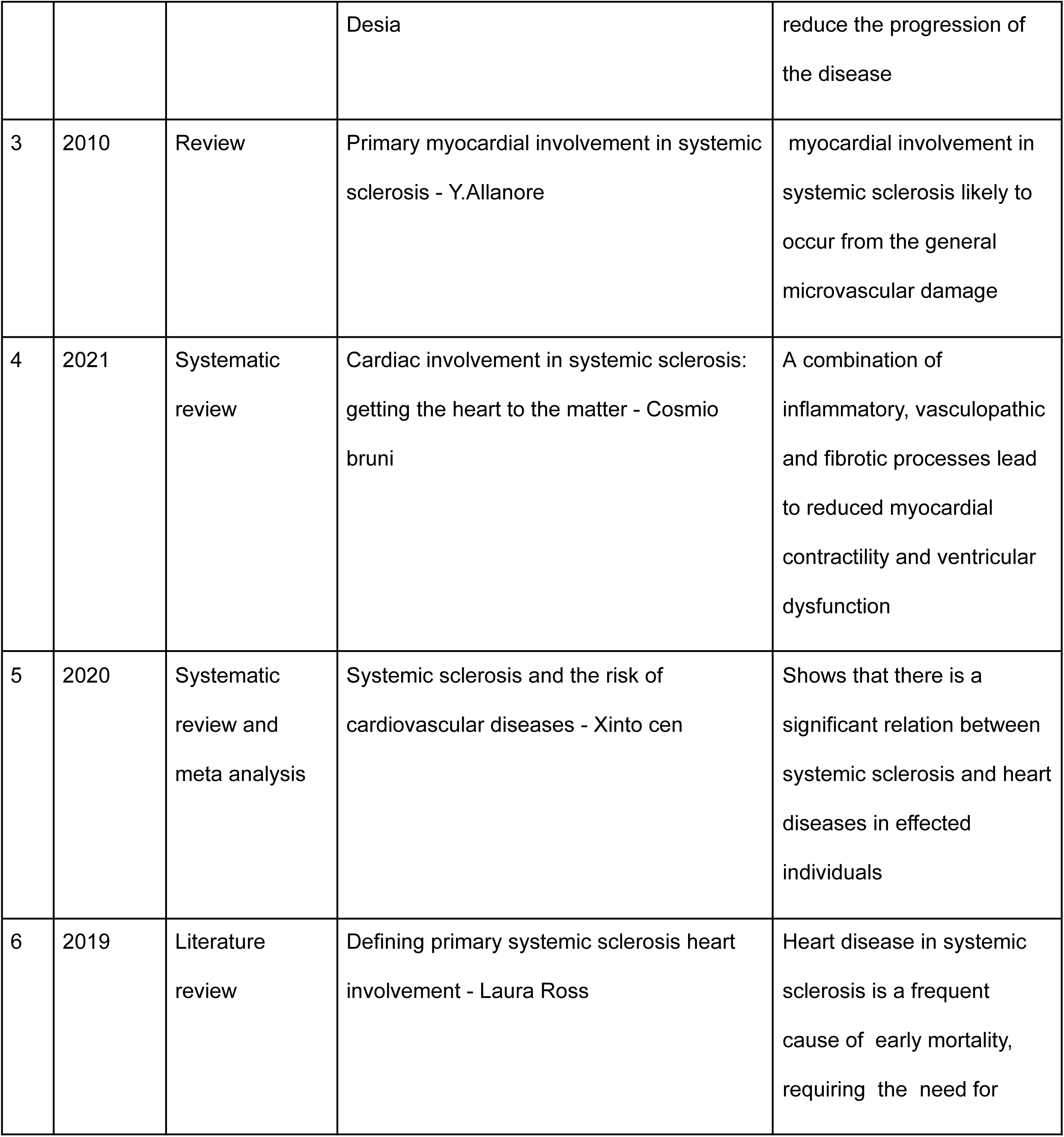
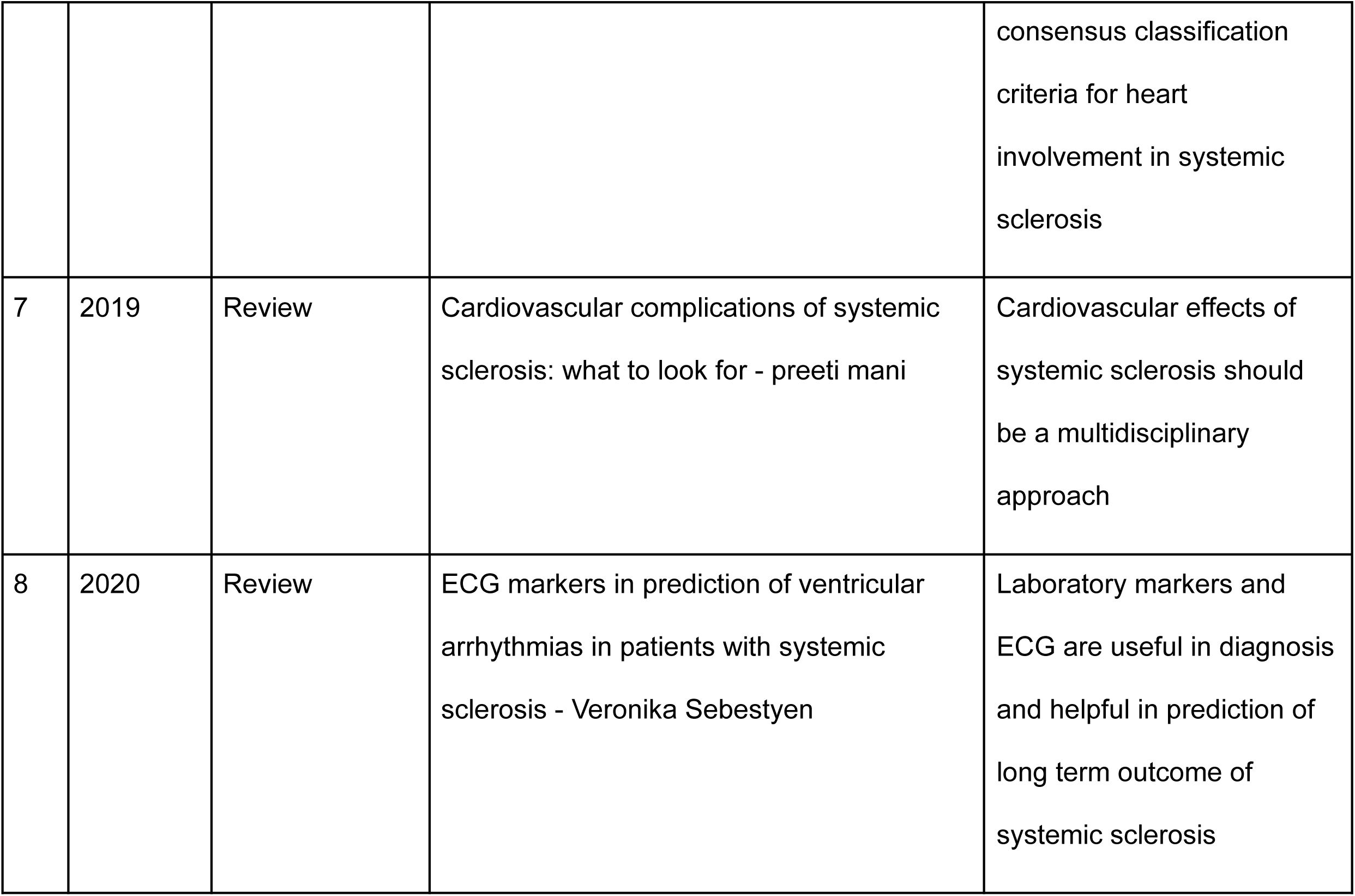
Summary of the results of the selected papers.

| **S.no** | **Year** | **Study design** | **Article name and Author** | **Conclusion** |
| --- | --- | --- | --- | --- |
| 1 | 2014 | Review | Cardiac arrhythmias and conduction defects in systemic sclerosis - Alexander Vacca | Arrhythmias and conduction defects are frequent manifestations of systemic sclerosis . They needed to be identified and treated early to improve the overall outcome of the disease |
| 2 | 2011 | Systematic review | Systemic sclerosis and the heart : current diagnosis and management - Chintan.s. | Vasodilators improve the myocardial perfusion and |
|  |  |  | Desia | reduce the progression of the disease |
| 3 | 2010 | Review | Primary myocardial involvement in systemic sclerosis - Y.Allanore | myocardial involvement in systemic sclerosis likely to occur from the general microvascular damage |
| 4 | 2021 | Systematic review | Cardiac involvement in systemic sclerosis: getting the heart to the matter - Cosmio bruni | A combination of inflammatory, vasculopathic and fibrotic processes lead to reduced myocardial contractility and ventricular dysfunction |
| 5 | 2020 | Systematic review and meta analysis | Systemic sclerosis and the risk of cardiovascular diseases - Xinto cen | Shows that there is a significant relation between systemic sclerosis and heart diseases in effected individuals |
| 6 | 2019 | Literature review | Defining primary systemic sclerosis heart involvement - Laura Ross | Heart disease in systemic sclerosis is a frequent cause of early mortality, requiring the need for |

TABLE 4: Summary of the results of the selected papers.
|  |  |  |  |  |
| --- | --- | --- | --- | --- |
|  |  |  |  | consensus classification<br>criteria for heart<br>involvement in systemic<br>sclerosis |
| 7 | 2019 | Review | Cardiovascular complications of systemic<br>sclerosis: what to look for - preeti mani | Cardiovascular effects of<br>systemic sclerosis should<br>be a multidisciplinary<br>approach |
| 8 | 2020 | Review | ECG markers in prediction of ventricular<br>arrhythmias in patients with systemic<br>sclerosis - Veronika Sebestyen | Laboratory markers and<br>ECG are useful in diagnosis<br>and helpful in prediction of<br>long term outcome of<br>systemic sclerosis |

## Discussion

Heart diseases in systemic sclerosis patients can be classified into 2 main abnormalities. The following items are a part of them.

### 1. Arrhythmias

Systemic sclerosis can cause electromechanical divergence and myocardial fibrosis leading to abnormalities in generation of pulse and conduction. As a consequence it can lead to a variety of arrhythmias but supra ventricular and ventricular arrhythmias have a significant place as these arrhythmias have a poor prognosis and can lead to sudden cardiac death. Arrhythmias are associated with poor prognosis and stands for 6% overall causes of death in the large European League Against Rheumatism (EULAR) Scleroderma Trials and Research (EUSTAR) database. From 128 death related to systemic sclerosis, 33 were cardiac related with almost half of them related to malignant arrhythmias. Report from the Genetics Versus Environment In Scleroderma Outcome Study (GENISOS) cohort, which includes 250 patients with a disease duration of 3 years, reported a total of 52 deaths related to systemic sclerosis. [1,3,10]

In a study by Ferri et al. [11], resting ECG shows some abnormal features in 22 out of 53(42%) SSc patients. Rhythm abnormalities were found in 30% of patients, but when 24-h Holter ECG monitoring was done, supraventricular arrhythmias were reported in 66% of systemic sclerosis patients and ventricular arrhythmias were seen in 90%, multiform ventricular premature beats in 40%, few ventricular tachycardia in 28%. Although ECG results were normal in half f the patients ventricular arrhythmias are detected in some patients with echocardiogenic abnormalities [3]. Malignant ventricular arrhythmias such as pulseless ventricular tachycardia and ventricular fibrillation, resulted in the third most common cause of death in systemic sclerosis patients, and are responsible for overall 5% mortality. [12]

Other than ventricular arrhythmias and dysfunction of sinus node, atrio-ventricular conduction delay can occur in 57% of the patients that occurs due to decrease in conduction velocity of abnormal atrial fibers. Resting membrane potential changes, action potential amplitude or maximal velocity of the action potential positive stroke, are all associated with depressed conduction velocity and abnormal excitability. [3]

Electrocardiography is the mainstay of investigation to detect arrhythmias. This is done if patients have symptoms like palpitations and continuous rhythm monitoring with holte or event monitoring should be done depending on the frequency of the symptoms [1]. Electrocardiography can be done as a screening test in patients with no abnormal symptoms once or twice a year. Q waves can occur in systemic sclerosis patients (especially those with diffuse cutaneous systemic sclerosis), notably in the precordial leads, even without any coronary artery disease symptoms.[1]

### 2. Conduction abnormalities

Conduction abnormalities can occur in 1/5*th* to 1/3rd of the patients with systemic sclerosis.[1,13,11]. The most common conduction abnormalities are left bundle branch block and atrioventricular block.[1,3]. Though conduction abnormalities are more common in systemic sclerosis patients with myocardial disease, specific abnormalities of the conduction system have not been the cause of death in most of the patients. Indeed, the conduction system appeared to be relatively preserved from the myocardial changes of systemic sclerosis, and the high incidence of conduction disturbances may be a result of particular damage to the specialized conduction tissue proximal to it. [14,3].

### 3. Coronary artery disease

CAD involvement is the result of alterations in the microvascular system, overproduction of collagen by altered fibroblasts resulting in extracellular matrix deposition and complex immune system dysregulation. A different and particular combination of the above mentioned mechanisms leads to inflammatory, ischemic and fibrotic lesions involving all the cardiac structures, including the conduction system. However, it is unclear that fibrosis is the only underlying pathology responsible for these conduction abnormalities. [5]

Man et al 38 described that the incidence of myocardial infarction in patients with systemic sclerosis was 4.4/1,000 persons/year, and the incidence of stroke was 4.8 /1,000 persons/ year, compared with 2.5 / 1,000 persons/ year for both myocardial infarction and stroke in healthy controls matched for age, sex, and time of entry.[15]

Indeed, evidence comes from histopathological studies which showed diffuse patchy fibrosis, with contraction band necrosis that is not related to epicardial coronary artery stenosis [16, 5], whereas other studies have revealed concentric intimal hypertrophy associated with fibrinoid necrosis of intramural coronary arteries [17]. Moreover, angina pectoris and myocardial infarction have been observed in SSc patients whose epicardial coronary arteries were normal.[5]

SPECT and Cardiac Magnetic resonance imaging (MRI) can be used for investigation of microvascular lesions of heart. Single photon emission computed tomography (SPECT), allowing the assessment of myocardial perfusion, has demonstrated evidence of reversible ischaemia together with irreversible lesions and demonstrated by the induction of coronary vasospasm by cold pressor provocation. Diagnosis is established by contrasting stress images with a additional set of images taken during at rest. This technique allows to distinguish diffuse patchy defects evocative of microcirculatory impairment to homogeneous large fixation defects related to coronary distribution suggestive of epicardial stenosis [5,8,9].

Brain Natriuretic Peptide(BNP) and N-Terminal-pro B-type Natriuretic Peptide(NT-pro-BNP) can raise in the setting of RV dysfunction, left ventricular systolic as well as diastolic dysfunction, pulmonary arterial hypertension and myocardial ischemia making them biomarkers for diagnosis of cardiac dysfunction. Some authors additionally found that an NT-proBNP cut-off of125 pg/ml was optimal for the detection of cardiac involvement in SSc.[4]

### Other cardiac lesions

Other cardiac lesions include pericardial effusion and valvular lesions. Pericardial effusions can be exudative in nature with a Lactate dehydrogenase of > 200 U/L. [20]. These effusions can be further divided into small and large lesions of which large lesions can develop even before the diagnosis of systemic sclerosis. Pericardial disease is evident clinically in 5% to 16% of patients with systemic sclerosis[18]; limited cutaneous systemic sclerosis patients have more pericardial disease than those with diffuse cutaneous type (30% vs 16%).[19,1]. Pericardial effusions can be diagnosed with echocardiography although small pericardial effusions can not be seen in echocardiography they can rarely lead to cardiac tamponade which is a fatal state.[1]

Valvular lesions in the systemic sclerosis patient population have a limited significance as these lesions have less prevalence when compared to the general population. Frequent mitral valve prolapse and mild mitral regurgitation is seen in patients with limited cutaneous systemic sclerosis.[1,21,22]

## Conclusion

Cardiac lesions in patients with systemic sclerosis should be identified early to optimize the quality of life of these patients. Signs and symptoms for volume overload, markers like BNP and NT-pro BNP, ECG, cardiac MRI, SPECT, echocardiography can be used for diagnosis of the early cardiac lesions to decrease the mortality. Early identification of cardiac lesions can be done with collaboration of cardiologists and rheumatology. Despite the recent advances in the diagnosis of the cardiac lesions some questions like algorithms for management, screening techniques, gold standard investigations for both diagnosis and screening remain unclear.

## Data Availability

ALL DATA PRODUCED IN THE PRESENT WORK ARE CONTAINED IN THE MANUSCRIPT

